# Perceptions and Attitudes Towards COVID-19 Vaccination Amongst Pregnant and Postpartum Individuals

**DOI:** 10.1101/2021.12.17.21267997

**Authors:** Molly R. Siegel, Mario I. Lumbreras-Marquez, Kaitlyn James, Brandon R. McBay, Kathryn J. Gray, Julianna Schantz-Dunn, Khady Diouf, Ilona T. Goldfarb

## Abstract

**Introduction:** This study aims to characterize attitudes towards COVID-19 vaccination and to evaluate factors associated with vaccine uptake amongst pregnant individuals.

**Methods:** An anonymous survey was distributed to a convenience sample of pregnant individuals receiving prenatal care at two large urban academic hospitals in a single healthcare network in Massachusetts. Individual demographic variables were included in the survey along with questions assessing attitudes towards COVID-19 and vaccination in pregnancy.

**Results:** Of 477 respondents, 233 (49.3%) had received or were scheduled to receive a COVID-19 vaccine. Age, White race, non-Hispanic/LatinX ethnicity, working from home, and typical receipt of the influenza vaccine were associated with COVID-19 vaccination. 276 respondents (58.4%) reported that their provider recommended the COVID-19 vaccine in pregnancy; these participants were more likely to have received a vaccine (OR 5.82, 95% confidence interval [CI] 3.68-9.26). Vaccinated individuals were less likely to be worried about the effects of the vaccine on themselves (OR 0.18, 95% CI 0.12-0.27) or their developing babies (OR 0.17, 95% CI 0.11-0.26). Unvaccinated individuals were less likely to report that it is easy to schedule a COVID-19 vaccine (OR 0.56, 95% CI 0.34-0.93), to travel to receive a vaccine (OR 0.19, 95% CI 0.10-0.36), and to miss work to receive a vaccine (OR 0.30, 95% CI 0.18-0.48).

**Conclusions:** Strategies are needed to improve patient education regarding vaccine side effects and safety in pregnancy and to change policy to make it feasible for pregnant patients to schedule and miss work without loss of pay to get vaccinated.

**Key Points:**

1. There were racial and ethnic disparities in COVID-19 vaccination.
2. Unvaccinated respondents were more likely to be concerned about vaccine effects for themselves or their growing babies.
3. Unvaccinated respondents cited work and scheduling-related barriers to vaccination, indicating areas for advocacy

## Introduction

Pregnant individuals are at increased risk for severe COVID-19, with higher rates of hospitalization, intensive care unit (ICU) admission, and death compared to their non-pregnant peers.^1-5^ COVID-19 is also associated with increased rates of adverse pregnancy outcomes such as pre-eclampsia, preterm birth, and cesarean delivery.^6-10^ Immunization with the messenger RNA (mRNA) and the viral vector COVID-19 vaccines has been shown to significantly reduce SARS-CoV-2 related morbidity and mortality, making immunizations essential to pandemic control. Unfortunately, pregnant individuals were not included in the initial COVID-19 vaccine trials, leading some providers to be reticent to recommend COVID-19 vaccination early in the vaccine rollout.^11,12^ However, recent post-approval studies suggest that COVID-19 vaccination is safe in pregnant and postpartum persons and effective in reducing maternal infections, morbidity, and mortality.^13-16^

In light of the post-authorization data demonstrating safety in pregnancy, the increased spread of new SARS-CoV-2 variants of concern, and the growing body of evidence surrounding the risks of COVID-19 in pregnancy, the American College of Obstetrics and Gynecology (ACOG), the Society for Maternal Fetal Medicine (SMFM), and the Centers for Disease Control and Prevention (CDC) now recommend COVID-19 vaccination in pregnancy.^17-19^ Despite this recommendation, pregnant persons continue to have lower vaccination rates than the general population.^20^ In the United States, COVID-19 vaccination in pregnant individuals remains lower than average rates of influenza and Tdap immunization, indicating that there are individuals who may typically accept vaccines who have not yet been vaccinated against COVID-19.^21^ Furthermore, globally observed racial and ethnic disparities in COVID-19 vaccination, infection, morbidity, and mortality persist in pregnant individuals and parallel that of the general population.^20,22^ Understanding factors associated with vaccine hesitancy is thus vital to improve vaccination rates, curb infection-related morbidity and mortality, and reduce COVID-19 related disparities.

Lack of inclusion of pregnant women in vaccine trials as well as lack of long-term safety data have been posited as reasons for low vaccine uptake in pregnancy.^23-25^ However, factors associated with COVID-19 vaccine acceptance in pregnancy remain understudied. In this study, we surveyed pregnant and postpartum individuals in the outpatient setting to characterize perceptions and attitudes towards COVID-19 vaccination and to evaluate factors associated with vaccine uptake and vaccine hesitancy.

## Methods

### Participants

Survey participants were from a cross-sectional convenience sample of pregnant and postpartum individuals aged 18 or older receiving prenatal care at two large urban academic hospitals and three affiliated community health centers in a single healthcare network in Massachusetts from June to August 2021. Participants were approached by study staff during outpatient prenatal and ultrasound visits and were offered participation. Consent to participate in the survey was implied by voluntary participation in the anonymous survey. The study and survey instrument were approved by the Mass General Brigham IRB (protocol # 2021P001086).

### Survey

A paper-based anonymous survey was designed to accommodate quantitative analysis of questions with nominal, ordinal, and interval level measurements. A copy of the survey is shown in the Appendix. Individual demographics were included in the survey along with questions assessing attitudes towards general vaccination, COVID-19 vaccination, and COVID-19 in pregnancy. The study was offered to participants in English and Spanish.

### Data Analysis

Data are presented as mean (standard deviation [SD]), median (interquartile range [IQR]), or number (%). For quantitative variables, descriptive statistics are presented to illustrate the distribution of the respondent demographics and survey responses. Associational odds ratios (ORs) were calculated via univariable logistic regression. Parametric or nonparametric tests (chi-square or Fisher’s exact test) were used to analyze categorical variables, after evaluation for cell size. Student’s *t*-test or Wilcoxon rank-sum tests were used to analyze continuous variables. A two-sided *P* <0.05 was considered statistically significant with correction for multiple testing where appropriate. Data were analyzed using Stata (IC 16.1, Stata Corp, TX).

## Results

### Patient Demographics

Between June-August 2021, 477 pregnant and postpartum individuals completed the survey out of 684 distributed, a response rate of 69.7%. 4 individuals did not report their vaccination status and were excluded from these analyses for a total of 473 responses in the final analyses: 453 were pregnant, and 19 postpartum with 1 respondent who did not report their pregnancy status. Overall, 233 individuals (49.3%) had received or were scheduled to receive a COVID-19 vaccine. Participant demographics by vaccination status are shown in Table 1. Age, working from home, and typical receipt of the influenza vaccine were associated with COVID-19 vaccination. Individuals who identified as Hispanic/LatinX were less likely to be vaccinated than those who identified as non-Hispanic/Latinx (37.3% of Hispanic/LatinX vs 55.4% of non-Hispanic/LatinX, p=0.003 after adjustment for multiple comparisons). Respondents who identified as Black or African American were less likely than those who identified as White to be vaccinated (18.3% of Black or African American vs 62.6% of White respondents, p<0.001). Additionally, those who identified their race as Other or whose race was Unknown were less likely than those who identified as White to be unvaccinated (32.4% vs 62.6%, p<0.001 for Other vs White and 32.8% vs 62.6%, p<0.001 for Unknown vs White). There were no differences in vaccination rates between pregnant and postpartum individuals (50.1% of pregnant respondents vs 30.6% postpartum, p=0.16).

**Table 1:** Demographic Characteristics by Vaccination Status.

|  | Vaccinated<br>(n=233) | Unvaccinated<br>(n=240) | P-value |
| --- | --- | --- | --- |
| **Age, mean y (SD)** | 33.0 (4.5) | 31.4 (5.6) | **<0.005** |
| **Ethnicity, n (%)** |  |  | **0.002** |
| Non-Hispanic/Latinx<br>(n=285) | 158 (67.8%) | 127 (52.9%) |  |
| Hispanic/Latinx (n=134) | 50 (21.5%) | 84 (35.0%) |  |
| Unknown (n=54) | 25 (10.7%) | 29 (12.1%) |  |
| **Race, n (%)** |  |  | **<0.005** |
| American Indian or<br>Alaska Native (n=3,<br>0.63%) | 0 (0%) | 3 (1.3%) |  |
| Asian (n=36, 7.6%) | 17 (7.3%) | 19 (7.9%) |  |
| Black or African<br>American (n=60, 12.7%) | 11 (4.7%) | 49 (20.4%) |  |
| Native Hawaiian or<br>Pacific Islander (n=1,<br>0.2%) | 1 (0.4%) | 0 (0%) |  |
| White (n=275, 58.1%) | 172 (73.8%) | 103 (42.9%) |  |
| Other (n=37, 7.8%) | 12 (5.2%) | 25 (10.4%) |  |
| Unknown (n=61, 12.9%) | 20 (8.6%) | 41 (17.1%) |  |
| **Number of People in Household, mean (SD)** | 2.96 (1.3) | 3.20 (1.5) | 0.07 |
| **Work Location n (%)** |  |  | **<0.005** |
| Working remotely<br>(n=139) | 87 (37.3%) | 52 (21.7%) |  |
| Working in person<br>(n=145) | 67 (28.8%) | 78 (32.5%) |  |
| Working both remotely<br>and in person (n=68) | 40 (17.2%) | 28 (11.7%) |  |
| Not currently working<br>(n=115) | 38 (16.3%) | 77 (32.1%) |  |
| Unknown (n=6) | 1 (.43%) | 5 (2.1%) |  |
| **Any medical comorbidities, n (%) \*** | 50 (21.5%) | 47 (19.6%) | 0.66 |
| **Typically receive influenza vaccination, n (%)** | 201 (86.2%) | 149 (62.1%) | **<0.005** |

\*Medical comorbidities were defined as self-report of cancer, renal disease, asthma, obesity, heart disease, hypertension, immune compromise, diabetes, sickle cell disease

225 of the vaccinated respondents (96.5%) disclosed the type of vaccine they received: 214 (95.1%) had received one or both doses of an mRNA vaccine (either Pfizer or Moderna), 10 (4.4%) had received a J&J/Janssen viral vector vaccine, and 1 (0.4%) was unsure of their vaccine type. Of unvaccinated individuals (n=240), 14 (5.8%) planned to receive a vaccine during pregnancy. 276 respondents (58.4%) agreed or strongly agreed that their provider recommended the COVID-19 vaccine in pregnancy; these participants were more likely to have received a vaccine (OR 5.82, 95% confidence interval [CI] 3.68-9.26).

### SARS-CoV-2 Infection Attitudes

Attitudes towards SARS-CoV2 infection between those who were unvaccinated and those who were vaccinated are shown in Figure 1. Vaccinated individuals were more likely than unvaccinated to agree or strongly agree that they feared COVID-19 during pregnancy, that if they were infected they were at risk for getting very sick, and that they knew someone who was hospitalized due to COVID-19. Those who were unvaccinated were more likely to agree or strongly agree that at some point in the pregnancy they believed they had SARS-CoV-2 infection and were more likely to have had a confirmed diagnosis of SARS-CoV-2 infection in pregnancy. One person in the cohort had been hospitalized due to COVID-19; that individual was not vaccinated.

**Figure 1:**
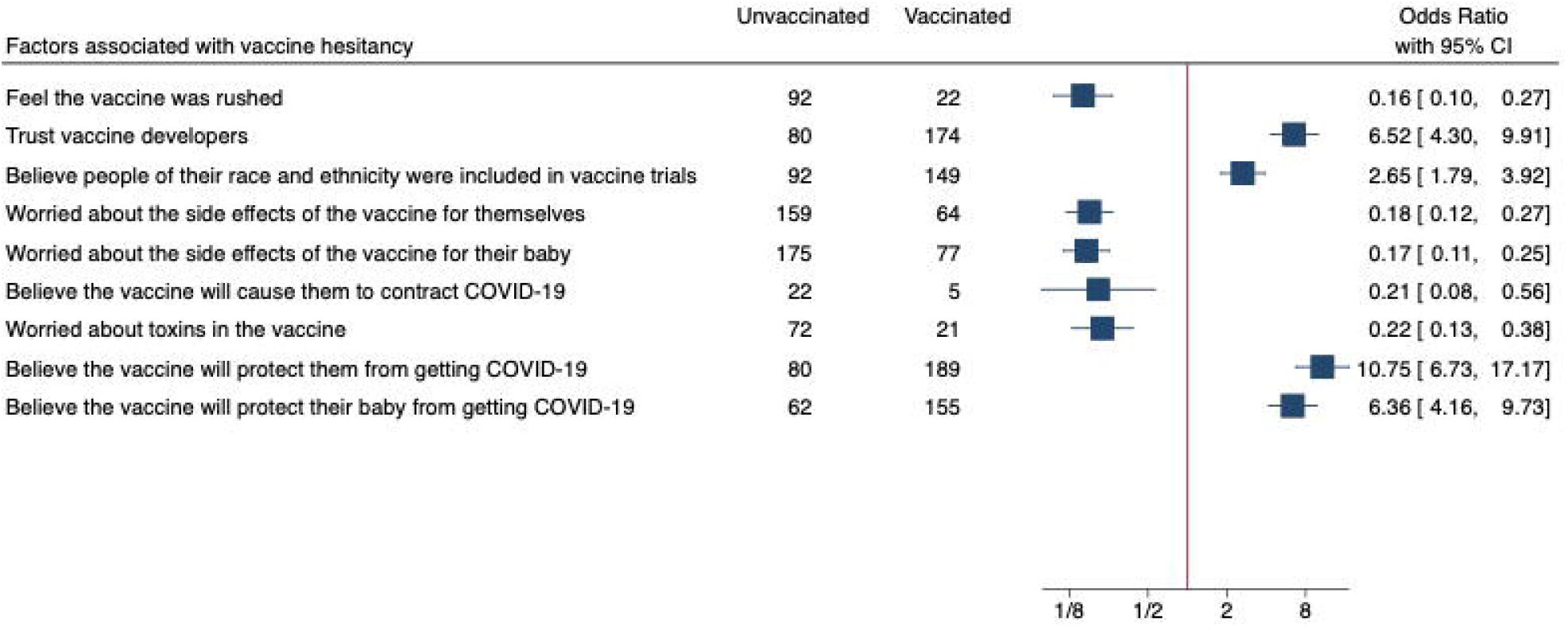
Attitudes Towards SARS-CoV2. *OR is relative to unvaccinated

### Factors associated with Vaccine Hesitancy

Factors associated with vaccine hesitancy are shown in Figure 2. Unvaccinated individuals were more likely to agree or strongly agree that the vaccine was rushed and that they were worried about toxins in the vaccine. They were also more likely to agree or strongly agree that they were worried about the side effects of the vaccine for themselves or their baby or that the vaccine would cause them to contract COVID-19. Unvaccinated individuals were less likely to agree or strongly agree that they trust vaccine developers and that people of their race and ethnicity were included in vaccine trials. Vaccinated individuals were more likely to agree or strongly agree that the vaccine would protect them or their baby from becoming infected.

**Figure 2:**
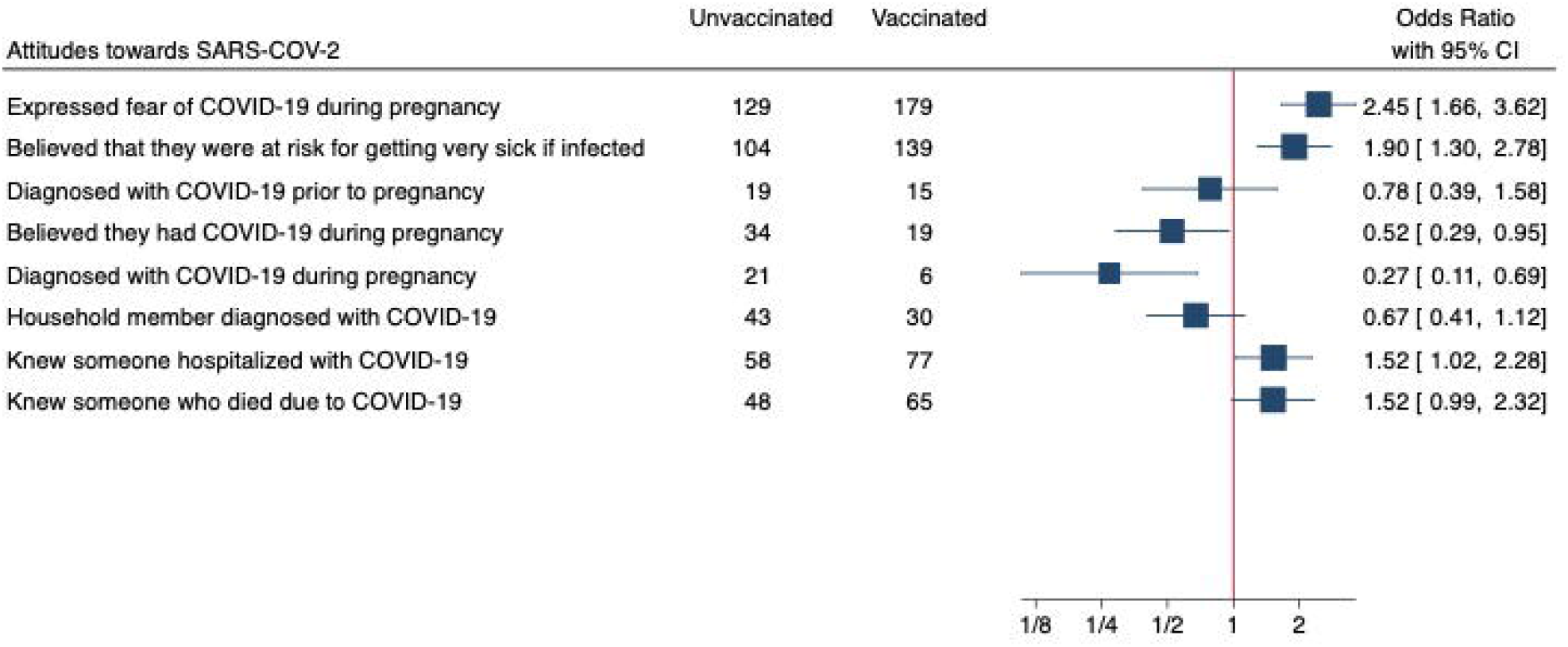
Factors Associated with Vaccine Hesitancy. *OR is relative to unvaccinated

### Barriers to Vaccination

Unvaccinated individuals were less likely to agree or strongly agree that it is easy to schedule a COVID-19 vaccine (OR 0.56, 95% CI 0.34-0.93), that it would be easy to travel to receive a vaccine (OR 0.19, 95% CI 0.10-0.36), that it would be easy to miss work to receive a vaccine (OR 0.30, 95% CI 0.18-0.48), and that it would be easy to miss time with family to receive a vaccine (OR 0.27, 95% CI 0.16-0.43).

### Effect of ACOG and CDC Statements

There were 35 respondents who completed the survey after the July 30, 2021 ACOG statement recommending COVID-19 vaccination in pregnancy and 23 who completed after the August 11, 2021 CDC statement recommending COVID-19 vaccination in pregnancy. There were no differences in self-reported vaccination rates in this cohort before and after the ACOG statement (p=0.98) or before and after the CDC statement (p=0.98). There were no changes to the above reported differences in baseline demographics, attitudes towards SARS-CoV2 infection, vaccine hesitancy, or barriers to vaccination after excluding those who responded after the ACOG statement (data not presented).

## Discussion

In this cross-sectional study, only half of pregnant women reported receiving at least one dose of a COVID-19 vaccine, and few of those who had not been vaccinated planned to receive a vaccine during pregnancy. While this number is higher than the global and United States national reported rates of COVID-19 vaccination in pregnancy, it is lower than the Massachusetts adult vaccination rate of 63.6% at the time of this survey distribution and lower than this group’s self-reported acceptance of influenza vaccination (74.7%).^20,26^ These findings add to the growing body of evidence pointing towards suboptimal COVID-19 vaccination rates in pregnant individuals. Our findings are similar to the survey results by Sutton et al. in New York City, which showed that pregnant respondents were less likely to accept vaccination compared with non-pregnant female respondents.^25^ Similarly, Turocy et al. performed a survey of patients seeking fertility treatment and found that those who were planning pregnancy or currently pregnant were less likely to accept vaccination than those who were not planning pregnancy.^27^ This may be because there is more vaccine hesitancy amongst pregnant individuals than non-pregnant individuals for vaccination in general. For example, prior studies have demonstrated suboptimal rates of influenza vaccination and Tdap vaccination in pregnancy, citing concerns about vaccine safety, lack of counseling, and disbelief in severity of disease.^21,28^

Our study identified several factors associated with receipt of COVID-19 vaccination. Those who worked remotely were more likely to be vaccinated, whereas those who were not currently working were less likely to be vaccinated. Those who were vaccinated against COVID-19 were more likely to report that they typically receive the influenza vaccine, indicating that in general they are more accepting of vaccination.

Importantly, there were racial and ethnic disparities in COVID-19 vaccination rates in this population. Only 42% of those who identified as Hispanic/LatinX and 19% of those who identified as Black and 32.4% of those who identified as Other reported receiving a vaccine, as compared with 55% of those who identified as non-Hispanic/LatinX and 64% of those who identified as White. Furthermore, those who had not received a vaccine were less likely to agree that people of their race and ethnicity were included in vaccine trials. These data mirror national trends in vaccination rates in non-pregnant individuals.^4^ These findings highlight a need for broader inclusion of individuals of non-White race in vaccine trials and vaccination campaigns to ensure equitable access to and trust in vaccination. As has been previously argued, the burden of these disparities in vaccination rates should not fall on the patients but rather on the health systems and providers that initiate and preserve inequity and mistrust.^29^

Survey respondents who had not received a COVID-19 vaccine were less likely to agree that they trusted vaccine developers and were more likely to feel that the vaccine was rushed. They were more worried about vaccine side effects and the effects of the vaccine on their developing fetus. These findings support the work by Skjefte et al., an international survey distributed electronically that found that pregnant individuals were worried about fetal effects, lack of safety data in pregnancy, and rushed development of vaccines.^24^ Our study builds on these data by comparing concerns between pregnant persons who are and are not vaccinated. Unvaccinated individuals were more likely to have been infected by SARS-CoV-2 during pregnancy and less likely to be fearful of COVID-19 or to believe that if they contracted SARS-CoV-2 they were at risk of becoming very sick. These findings provide areas for improved patient counseling surrounding the risks of SARS-CoV-2 infection in pregnancy and the safety of vaccination, as post-approval studies have not shown more severe side effects in pregnant individuals or detrimental effects of the vaccine on pregnancies or fetal and neonatal outcomes.^30,31^ They also highlight the importance of ongoing long-term follow-up studies to evaluate the health of children exposed to vaccination during pregnancy to provide families with long-term safety data.

Respondents who were not vaccinated against COVID-19 identified several barriers to vaccination, including concerns about missing work or time with family as well as difficulty scheduling a vaccine. These are thus areas for advocacy to diminish the effects of these logistical barriers: vaccine recovery days could be protected and paid time off so that patients do not worry about missing time from work, vaccines can be mandated at workplaces to normalize vaccination and missing work to attend vaccine appointments, and they should be made available at expanded hours and at places that are frequently visited such as at workplaces and grocery stores.^32-34^ Reducing some of these barriers to vaccination may lead to higher vaccination rates and reduced inequities in immunization status observed in our study and previous analyses.

While most respondents in this study had discussed COVID-19 vaccination with their obstetric providers, just over half reported that their provider recommended COVID-19 vaccination in pregnancy. Given that provider confidence and recommendation of vaccination has consistently been shown to be a strong predictor of patient vaccination rates particularly in a pregnant population, it is imperative that providers feel comfortable engaging in conversations with patients surrounding immunization.^35,36^ Now that the CDC and ACOG both recommend COVID-19 vaccination in pregnancy, providers should routinely discuss and recommend these vaccines to their patients with attention to addressing myths, misconceptions, and logistical barriers. National and institution-specific counseling guides may assist in the implementation of these recommendations.

Strengths of this analysis include its large sample size and high response rate as well as diverse patient population. It is novel in its assessment of barriers to vaccination and comparisons of vaccination concerns amongst pregnant individuals who were and were not vaccinated. It was also conducted when all patients were eligible to receive vaccination, thus eliminating any hesitation due to lack of eligibility. Limitations include that it was conducted when the CDC and ACOG guidance was to offer, but not necessarily recommend, vaccination to pregnant patients. Acceptance of vaccination may be higher now that these organizations have provided stronger guidance regarding vaccination in pregnancy. Furthermore, the survey did not ask participants to differentiate their responses based on receipt or consideration of mRNA or viral vector vaccine, and it is possible that participants may have had different concerns depending on vaccine type.

## Conclusions

Given the known increased risks of SARS-CoV-2 infection in pregnancy and the ongoing high case rates globally, vaccination of pregnant individuals is paramount to halting disease transmission and reducing barriers in SARS-CoV-2-related morbidity and mortality. An understanding of the factors associated with reduced vaccine uptake and the socioeconomic barriers to vaccination enables action to improve patient counseling and eliminate disparities in vaccination rates amongst pregnant individuals.

## Data Availability

All data produced in the present study are available upon reasonable request to the authors

## Acknowledgements

Dr. Gray has consulted for Illumina Inc,, Aetion, and BillionToOne outside the scope of the submitted work. No other financial disclosures are reported by the authors of this paper.

